# Race and ethnicity do not impact eligibility for remdesivir- a single-center experience

**DOI:** 10.1101/2020.12.29.20249004

**Authors:** Lauren Pischel, Makeda Walelo, Jemma Benson, Rebecca Osborn, Rachel Schrier, Jessica Tuan, Lydia Barakat, Onyema Ogbuagu

**Author notes:** Corresponding author: (LP).

## Abstract

As the Coronavirus-2019 (COVID-19) pandemic continues, multiple therapies are rapidly being tested for efficacy in clinical trials. Clinical trials should be racially and ethnically representative of the population that will eventually benefit from these medications. There are multiple potential barriers to racial and ethnic minority enrollment in clinical trials, one of which could be that inclusion and exclusion criteria select for certain racial or ethnic groups disproportionately. In this observational cohort study at a single health care system, we examined if there were differences in eligibility for treatment with remdesivir based on clinical trial criteria for racial and ethnic minorities compared to non-Hispanic Whites. 201 electronic medical record charts were reviewed manually. Self-identified Whites were older than other racial or ethnic groups. At the time of presentation, Black, Latinx, and White participants met inclusion criteria for remdesivir at similar rates (72%, 80%, and 73% respectively), exclusion criteria at similar rates (43%, 38% and 49% for Black, Latinx and White participants respectively). In this study, there was no difference in eligibility for remdesivir based on race or ethnicity alone.

## Introduction

As the Coronavirus-2019 (COVID-19) pandemic continues to spread across the United States, marked disparities in outcomes have emerged among racial and ethnic minorities. Cases of COVID-19 are not only higher in minorities but so are rates of hospitalization and death (1-3). While the mortality rate for White individuals with COVID-19 is about 54 per 100,000 people, for Black individuals it is two-fold higher at 108 per 100,000, while the mortality rate for Latinx individuals is 74 per 100,000 and is 90 per 100,000 for indigenous individuals (3).

The cause for this racial and ethnic disparity is not rooted in genetics, but rather from a series of interlinked factors including structural racism, long-standing healthcare inequities, a greater prevalence of underlying co-morbidities, and participating in essential jobs leading to increased risk of exposure to the virus (4-10). Indeed, COVID-19 has exacerbated the already existing health inequity in the United States (11).

Historically, racial and ethnic minorities have been under-represented in clinical trials (12). The reasons for this are multiple, including distrust arising from prior injustices dating back decades, hidden costs of clinical trials, lack of accessibility to clinical trial sites, lack of prior community participation, and bias from researchers (13, 14). The lack of a broad representation in clinical trials is not only an issue of justice but also represents a scientific problem as it impacts the external validity of clinical trial results which form the basis of ‘high quality’ evidence in medicine.

The SIMPLE studies evaluated the anti-viral remdesivir as a treatment for COVID-19 and these were some of the earliest treatment trials launched during the first wave of the pandemic in the United States. This drug has since been shown to decrease time to clinical improvement in patients with moderate and severe COVID-19 (15). The SIMPLE studies and other clinical trials for therapeutics remain vitally important in determining which treatments have favorable impact on morbidity and/or mortality. However, eligibility criteria for therapeutic trials are not always inclusive of all patients with COVID-19 and thus may exclude individuals with certain characteristics, therefore limiting evaluation of therapies in those individuals and their subsequent ability to experience any therapeutic benefit.

In this observational cohort study of patients admitted to an academic healthcare system that serves a diverse patient population, we sought to examine if there were differences in eligibility for treatment with remdesivir for racial and ethnic minorities compared to non-Hispanic Whites, based on the eligibility criteria for the clinical trial, thus impacting their candidacy for a disease modifying and potentially life-saving treatment for COVID-19.

## Materials and Methods

### Study location

Data was obtained from patients hospitalized at Yale-New Haven Hospital (YNHH) from March 13^th^, 2020 through May 6^th^, 2020. YNHH has a 1,540 bed capacity and is the second largest hospital in the United States (16). The health system provides medical services to the greater New Haven area, a location that has a diverse patient population base with about a third of the population being Black, White or Hispanic and or Latinx respectively (17).

### Study population

All adult patients (ages 18 years or older) who were hospitalized with a positive SARS CoV-2 PCR test were eligible for medical record review. Subjects with missing race and ethnicity information were excluded from eligibility criteria analyses.

### Data collection

Data were primarily manually extracted by study investigators from patients’ electronic medical record (EPIC©, Epic Systems Corporation, Server Version May 2020, Verona, Wisconsin, USA) though some baseline laboratory values and admission times had been previously extracted via automated software. Each study investigator was assigned patients to review in blocks of 10 starting from the first COVID-19 patient admitted. Data was reviewed by another investigator, with discrepancies resolved by agreement. Demographic variables extracted included age, gender, race and ethnicity, insurance status, and body mass index (BMI). Race and ethnicity are entered by patients into their chart during clinical encounters. Patients who identified as Hispanic or Latino were noted as Latinx for this study. Clinical data extracted also included underlying medical conditions such as immunocompromised state (cancer treatment in the past year, use of immunosuppressive drugs, bone marrow or solid organ transplant recipient, HIV-positive status), duration of symptoms prior to diagnosis and hospital admission, vital signs, peripheral oxygen saturation, and disease severity defined by requirements for oxygen supplementation, vasopressors, mechanical ventilation, and intensive care unit (ICU) admission.

Laboratory values in the first 24 hours of hospital admission were captured, including complete blood counts, electrolyte profile, liver function tests, ferritin, lactate dehydrogenase (LDH), D-dimer, inflammatory markers, (C-Reactive Protein) cytokine profiles (interleukin-6), and radiologic studies where available. Oxygenation and oxygen support requirements in the first 24 hours were logged in utilizing the 7-category ordinal scale (18).

Patient clinical characteristics and laboratory tests in the first 24 hours of admission were matched to inclusion and exclusion criteria from remdesivir SIMPLE trials (which was the primary therapeutic clinical trial enrolling at this hospital during the study timeframe) (19). Where there were multiple measurements of a test or other variables within the 24-hour period, the most deranged value was recorded. All data was stored in a secure database with Microsoft Excel. De-identified data can be found at https://datadryad.org/stash.

### Ethical Considerations and Study Approval

The study was approved by the Yale University Human Investigation Committee. As this was a retrospective study with minimal risk, informed consent was not required or obtained.

### Analysis

For ease of analysis, patients were assigned first by their ethnicity and then by their race and categorized as follows: Hispanic/Latinx ethnicity of any race, Non-Hispanic Black, and Non-Hispanic White (subsequently referred to as White). Groups with less than five participants were excluded from comparative analysis. Descriptive statistics were conducted with continuous data presented as the mean with corresponding range and categorical data presented as absolute values and corresponding percentages. Continuous data were compared using one-way Analysis of variance (ANOVA) and categorical data were analyzed using Χ^2^ test or Fisher’s exact test. Post-hoc analysis for Χ^2^ for within group differences of baseline characteristics was assessed with the Χ^2^ test with Bonferroni correction as noted in the text. Group differences by race for inclusion and exclusion criteria were detected by a Χ^2^ test. *P-*values⍰less than⍰0.05 were considered statistically significant, and all reported *P*-values are two-tailed. Analysis was completed in JMP version 15 statistical software (SAS Institute, Cary, North Carolina) and R version 4.0.2 (R Core Team, Vienna, Austria.)

## Results

Of 1,591 adults admitted with COVID-19 during the study period, 203 patient charts were reviewed starting at the first patients admitted, some reviewers were able to complete more blocks of 10 than other reviewers so the 203 patients were not consecutive admissions. 2 excluded as they were less than 18 years of age. Of the remaining 201 patients, the mean age of patients was 62.8 years (21-99 years) but was significantly lower among Latinx participants (49.2 years compared to 68.7 years and 62.3 years for non-Hispanic Whites and Blacks respectively [*P* <0.0001]) (Table 1). A slight majority of study participants were men 106/201 (53%) but the gender distribution varied significantly between Black participants where majority were women (63%) while majority of Latinx and White patients were men (73 and 56% respectively [*P* =0.002]). Regarding underlying medical conditions, there were significant differences between groups as 67% of Black participants and 55% of White participants had hypertension while it was present in only 23% of Latinx individuals (P<0.001). Rates of diabetes were also significantly different between groups with a prevalence of 44% among Black participants which was significantly greater than the 15% and 22% in Latinx and White participants (P=0.002). Prevalence of chronic kidney disease was lower in Latinx patients (15%) compared with Black patients (37%) and White patients (27.6%), this difference approached but did not reach significance[*P*=0.060]. Otherwise, there were no significant differences between groups regarding the proportion of immunocompromised patients (13% overall), however, there were more HIV-positive patients who were Black (8%) and Latinx (3%) than White (none) [*P* =0.012]. The mean time from symptom-onset to having a positive COVID-19 test and hospital admission were similar between groups (mean of 4.8 days and 5.8 days respectively). Overall, 10% of patients were critically ill requiring vasopressor support on the day of admission.

**Table 1:** Demographics and Clinical Characteristics of the Study Cohort.

| Characteristic | Total Patients (n = 201) [% or range] | Black (n = 60) | Latinx (n = 40) | White (n = 98) | P-value |
| --- | --- | --- | --- | --- | --- |
| Age in years, mean | 62.8 (21-99) | 62.3 (23-93) | 49.2 (22-72) | 68.7 (21-99) | <0.0001* |
| Gender no. (%) |  |  |  |  | 0.0020* |
| Woman | 95 (47.2%) | 38 (63.3%) | 11 (27.5%) | 43 (43.9%) |  |
| Man | 106 (52.7%) | 22 (36.7%) | 29 (72.5%) | 55 (56.1%) |  |
| Body mass index (kg/m<sup>2</sup>) | 30.5 (16 - 59.6) | 33.0 (19.6-59.6) | 30.5 (16.0 - 45.8) | 29.2 (17.1-53.6) | 0.012* |
| Hypertension | 105 (51.7%) | 40 (66.7%) | 9 (22.5%) | 54 (55.1%) | <0.0001* |
| Diabetes | 54(26.8%) | 26 (43.3%) | 6 (15.0%) | 22(22.4%) | 0.0024* |
| Chronic Kidney Disease | 55 (27.4%) | 22 (36.7%) | 6 (15.0%) | 27 (27.6%) | 0.0602 |
| Immunocompromised | 26 (12.9%) | 12 (20%) | 3 (8%) | 10 (10%) | 0.1091 |
| Cancer treatment in the last year | 11 (5.5%) | 3 (5%) | 2 (5%) | 6 (6%) | 0.9423 |
| Use of immunosuppressive medications | 8 (4.0%) | 3 (5%) | 0 (0%) | 4 (4 %) | 0.3811 |
| Bone marrow or solid organ transplant recipient | 7 (3.5%) | 3 (5%) | 0 (0%) | 4 (4 %) | 0.3811 |
| HIV-positive status | 6 ( 3.0%) | 5 (8.3%) | 1 (2.5%) | 0 (0%) | 0.0120* |
| Hospital Length of Stay, days | 12.4 (1.3 - 96.0) | 10.1 ( 1.4 - 78.0) | 12.8 (1.8 - 96.0) | 13.3 (1.3 - 89.8) | 0.3369 |
| Time to Positive Test:<br>Mean difference<br>between symptom-onset<br>and COVID-19 positive<br>test (range of days) | 4.8 (0-22) | 4.9 (0 -22) | 5.7 (0-16) | 4.4 (0-19) | 0.4472 |
| Symptom duration: Mean<br>difference between<br>symptom-onset and date<br>of admission test (range<br>of days) | 5.8 (0 - 35) | 5.1 (0-22) | 6.9 (0-35) | 5.9 (0 - 33) | 0.3539 |
| Pressor requirement first<br>24 hours | 20 (10.0%) | 6 (10.0%) | 3 (7.5%) | 11 (11.2%) | 0.8046 |
| *: P value less than or<br>equal to 0.05 |  |  |  |  |  |

Examining the data by race, three participants were excluded including a single individual identifying as Asian and two individuals who did not have any specific race and ethnicity designations leaving 198 participants for the remainder of the analysis (Table 2).

**Table 2:** Study Participants and Eligibility Criteria for Remdesivir.

| **Number of people meeting**<br>**Inclusion criteria** | **Black (n = 60)** | **Latinx (n = 40)** | **White (n = 98)** | **P-value** |
| --- | --- | --- | --- | --- |
| ≥ 18 years old (or ≥ 12 years old ≥ 40 kg) | 100% | 100% | 100% |  |
| SARS-CoV-2 RNA<br>test confirmed positive ≤ 4<br>days before randomization | 100% | 100% | 100% |  |
| SpO2 ≤ 94% on<br>room air or requiring<br>supplemental oxygen at<br>screening (severe COVID-19<br>arm) | 71.7 % (n = 43) | 80.0% (n = 32) | 80.0% (n = 78) | 0.4623 |
| SpO2 > 94% on<br>room air at screening<br>(moderate arm) | 28.3% (n = 17) | 20.0% (n=8) | 20.4% (n = 20) | 0.4623 |
| Radiographic<br>evidence of pulmonary<br>infiltrates | 68.3% (n = 41) | 80.0% (n =32) | 73.4% (n = 73) | 0.4182 |
| Met all inclusion<br>criteria | 68.2% (n = 41) | 80.0% (n =32) | 74.5% (n = 73) | 0.342 |
| **Number of people meeting**<br>**Exclusion criteria** |  |  |  |  |
| Multi-organ failure<br>(in first 24 hours) | 0% | 0% | 1.0% (n = 2) | 0.3531 |
| Mechanically ventilated (including V-V ECMO) ≥ 5 days, or any duration of V-A ECMO | 0% | 0% | 0% | 1 |
| ALT or AST $>$ 5 x upper limit of normal | 16.7% (n = 10) | 25.0% (n = 10) | 14.4% (n = 14) | 0.3117 |
| Creatinine clearance $<$ 50 mL/min (Cockcroft-Gault equation) | 31.7% (n = 19) | 12.5% (n = 5) | 31.6% (n = 31) | 0.0567 |
| Positive pregnancy test | 0% (n = 0) | 2.5% (n = 1) | 5.1% (n = 5) | 0.0998 |
| Met 1 non-modifiable exclusion criteria | 43.3% (n = 26) | 37.5% (n = 15) | 49.0% (n = 48) | 0.4485 |
| Met Composite of all inclusion and exclusion criteria | 40% (n = 24) | 50% (n = 20) | 42.8%(n = 42) | 0.6056 |

When looking at inclusion criteria for the remdesivir clinical trial, 100% of all participants met age criteria, and there were no significant differences with regards to proportion of participants that met criteria for the severe COVID-19 trial – 72% for Black participants, 80% for Latinx participants, and 80% for White participants. There were also similar frequencies of patients exhibiting pulmonary infiltrates with 68.3% of Black, 80% of Latinx and 75% of White participants having those findings on radiographic studies. The overall proportion of individuals meeting inclusion criteria for either the moderate or severe arm of remdesivir was similar across the three groups (*P* = 0.342).

As for exclusion criteria for remdesivir, elevated transaminase levels (alanine aminotransferase (ALT) and aspartate aminotransferase (AST)) greater than five times the upper limit of normal were similar across the groups, at 16.7%, 25.0%, and 14.4% for Black, Latinx and White patients respectively (*P* = 0.312). Creatinine clearance less than 50 mL/min was present in 31.7% of Black participants and 31.6% of White participants, while only present in 12.5% of Latinx patients. This difference approached but was not statistically significant (*P* = 0.0567). Five White patients and one Latinx patient were pregnant on admission. Overall, 86 patients met inclusion criteria (43%) and the proportion of patients meeting eligibility criteria for remdesivir (all inclusion and exclusion criteria) were not different by race (*P* = 0.6056).

## Discussion

Clinical trial enrollment of historically underrepresented groups such as racial and ethnic minorities should be a priority for all clinical trials, but especially those involving COVID-19 therapeutics and vaccines due to the disproportionate impact of the disease on those populations (12, 20). We sought to evaluate whether race/ethnicity was associated with differential eligibility for pivotal remdesivir trials, the only approved SARS CoV-2 antiviral to date. In this single center study, while evaluating cases early in the COVID-19 pandemic, we did not find that Black, Latinx, or White patients met the inclusion or exclusion criteria for the remdesivir clinical trial at different proportions either by individual inclusion and exclusion criteria, or by a composite measure of the clinical trial criteria. Notably there were differences in patient demographics where White and Black participants were older than Latinx participants and more likely to have age-related comorbidities such as hypertension. Black participants were more likely to have diabetes. This analysis provides reassuring data that the clinical trial eligibility criteria were not differentially exclusive of any racial/ethnic group. In the SIMPLE studies, 19% of the study participants were Black in the 5-day remdesivir arm and 13% were Hispanic or Latino (19). Ensuring a diverse group of study participants is imperative for all medications in clinical trials, but this is particularly important during a pandemic when a large portion of the population may receive medication shortly after FDA approval.

Certain derangements in laboratory tests are important for remdesivir dosing considerations. The intravenous form of remdesivir contains sulfobutylether-beta-cyclodextrin (SBECD) that is renally excreted. Though SBECD has been shown to be relatively safe in humans, in animal studies, repeat dosing is associated with renal tubular vacuolation and foamy macrophage accumulation in the liver and lungs; therefore, it is best avoided in individuals with impaired renal function (21). This is an important issue as chronic kidney disease (CKD) tends to differentially occur amongst older individuals and Black individuals, otherwise key populations for COVID-19 therapeutics, thus lower glomerular filtration rates (GFR) would then limit their candidacy for the drug. This was evident in our study where White and Black participants who were older than Latinx individuals, had a higher baseline proportion of CKD and had higher numerically proportions of patients excluded based on the GFR threshold, which approached but did not meet statistical significance.

Significant elevation of transaminases is also a notable exclusion criteria for the use of remdesivir. This exclusion is based on the risk of liver injury with the drug that occurs in about 5% or more of patients who are treated with the drug and may be a reason for treatment discontinuation due to severe and symptomatic elevation (22, 23). However, elevated transaminases may occur due to other etiologies including COVID-19 itself, such as complications such as ischemic liver injury from shock, the immune response to the infection (cytokine storm), drug-induced liver injury, or due to underlying chronic liver diseases. The prevalence of chronic liver diseases, such as hepatitis B and C infections do differ by race (24). Though this eligibility criterion is not modifiable, it may differentially exclude patients with more severe forms of COVID-19 such as those who present later and with greater metabolic derangements. Though not demonstrated in our study, Black patients in larger cohorts have been shown to present with more severe disease(25).

This study has several notable limitations: first, our single center experience may not be reflective of settings with dissimilar population characteristics. Second, our study evaluated eligibility at the time of admission, but this could change as a patient’s clinical status evolves over the course of admission either in a positive or negative direction, so it may not be applicable to patients who require prolonged hospitalization. Third, data was derived from pre-existing medical records, such that accuracy and timing of documentation could have impacted categorization of patient’s characteristics vis a vis eligibility criterion. Fourth, this analysis only looked at one clinical trial screening criteria and so may not be generalizable to other COVID-19 therapeutic trials. Finally, and most importantly, this study was limited by small sample size and so could have missed potential associations and examining larger samples would be a fruitful area for potential research.

## Conclusion

To conclude, our study did not find any differences in eligibility for remdesivir SIMPLE studies by race and ethnicity based on patient’s admission. The most common non-modifiable exclusion criteria was a GFR less than 50ml/min. Older age may result in decreased eligibility for remdesivir due to higher proportion of participants with reduced GFR.

## Data Availability

De-identified data can be found at https://datadryad.org/stash (in process of being uploaded)

